# Prevention of Atopic Diseases with the Use of Hypoallergenic Infant Formula: A Systematic Review and Meta-Analysis

**DOI:** 10.64898/2026.01.26.26344870

**Authors:** Helena M Vind, Morten A Rasmussen, Ann-Marie M Schoos

## Abstract

**Background:** Atopic diseases are estimated to affect 30-40% of the global population. However, the potential protective effect of hypoallergenic infant formula against conditions such as atopic dermatitis (AD), cow’s milk protein allergy (CMPA), and asthma remains uncertain.

**Objective:** To conduct a systematic review and meta-analysis of randomized controlled trials (RCTs) evaluating hypoallergenic formula for atopic disease prevention in high-risk infants. The primary outcome was AD and secondary outcomes were CMPA and asthma.

**Methods:** A systematic review and meta-analysis was conducted according to PRISMA 2020. RCTs involving high-risk infants were identified through PubMed, Cochrane Library, and Web of Science. Exclusion criteria included interventions not initiated at birth, enrolment of sick infants, and non-RCTs. Pooled Relative Risks (RR) with 95% confidence intervals (CI) were calculated using a random-effects model.

**Results:** We included 9 RCTs that enrolled high-risk infants. The meta-analysis found a borderline significant protective effects of AD (RR=0.78 [0.59-1.03], p=0.059; I^2^=46.5%), a significant protective effect of hypoallergenic formula in prevention of CMPA (RR=0.51 [0.27-0.97], p=0.0228; I^2^=37.3%), and no significant risk reduction for asthma (RR=0.78 [0.51-1.20], p=0.059; I^2^=37.5%).

**Conclusion:** This systematic review and meta-analysis found no statistically significant protective effect of hypoallergenic formula for AD or asthma, though a non-significant trend toward risk reduction was observed. A significant risk reduction was seen for CMPA (RR≈0.5), although not all diagnoses were confirmed by oral food challenge. These findings suggest potential patient-specific benefits, but larger, well-designed RCTs are needed to confirm them.

## Introduction and background

Atopic diseases affect an estimated 30-40% of the global population(1). Susceptibility to atopic disease is partly inherited through genetic predisposition(2), yet it is also substantially modulated by early-life environmental exposures, i.e. risk is increased following cesarean section and reduced in children raised in farming environments(3). It remains uncertain if the use of hypoallergenic formula in infancy has an effect in preventing atopic diseases such as atopic dermatitis (AD), cow’s milk protein allergy (CMPA), and asthma. In this systematic review, atopy is defined as an innate, hereditary tendency to develop atopic conditions such as AD, asthma, allergic rhinitis, and food allergies. A systematic review from 2021 reported that 6-10% of infants globally are affected by food allergy(4) and up to 20% by AD (5). These conditions place a considerable burden on families as well as healthcare systems(6,7). Furthermore, the incidence is increasing and a potential lack of immune system training in early life could contribute to this(8).

Hypoallergenic Infant Formula (HIF) is produced by hydrolyzing cow’s milk proteins into smaller peptides and, in some cases, to amino-acids, thereby reducing allergenic epitopes in the formula. It is recommended for infants diagnosed with CMPA who are not exclusively breastfed(9).

Depending on the degree of protein-breakdown, HIFs are categorized as partially hydrolyzed (pHF), extensively hydrolyzed (eHF) or elemental formula (EF) also known as amino-acid based formula(9).

Infants diagnosed with CMPA who receive hypoallergenic formula are exposed to fewer allergenic proteins while maintaining adequate nutritional intake. In most cases, eHF is enough for symptom resolution, whereas EF is reserved for infants where symptoms persist despite intake of eHF(10).

In addition to its therapeutic use, hypoallergenic formula has historically been recommended for infants at increased risk of atopic disease, particularly when exclusive breastfeeding cannot be maintained or needs to be supplemented during the first week of life(10). This approach has been adopted in some neonatal departments and postdischarge feeding guidance for high-risk newborns - such as those with a strong family history of allergy (e.g., AD, asthma, or food allergy in parents or siblings). The rationale is that early exposure to partially or extensively hydrolyzed protein may reduce sensitization to intact cow’s milk proteins and thereby help prevent allergic manifestations later in infancy. However, evidence for this preventive strategy remains inconsistent, which underscores the need for systematic evaluation through meta-analysis.

This systematic review and meta-analysis aims to synthesize and compare evidence from randomized controlled trials (RCTs) evaluating the effect of hypoallergenic formula initiated from birth on the prevention of atopic diseases in high-risk infants. The primary outcome is the development of AD, while secondary outcomes include CMPA and asthma.

### Rationale and Objectives

Atopic diseases are complex, multifactorial conditions in which genetic susceptibility interacts with early-life environmental factors. Among these, the influence of early nutritional exposures remains insufficiently understood. The preventive use of hypoallergenic infant formula in early infancy is based on the hypothesis that reducing exposure to intact cow’s milk proteins may decrease sensitization and thereby help interrupt the so-called “atopic march” - the progressive development of allergic conditions such as AD, food allergy, and asthma.

## Methods

The review protocol was registered in PROSPERO (CRD420251131139) and conducted in accordance with PRISMA 2020.

A search was made in the biomedical literature-database *PubMed* using the following search-string: “*(hypoallergenic infant formula OR hydrolyzed formula OR amino acid based formula) AND (allergy prevention OR allergic disease OR food allergy OR atopic dermatitis OR asthma) AND (infant OR newborn OR baby) AND (“randomized controlled trial”[Publication Type])*”.

In *The Cochrane Library*, the following search was used: (“partially hydrolyzed formula” OR “extensively hydrolyzed formula”) AND (“atopic dermatitis” OR “allergy”) AND (infant OR baby)

Lastly, in Web of Science, the following search was used: *TS = (“hypoallergenic infant formula” OR “hydrolyzed formula” OR “amino acid based formula”) AND TS = (“allergy prevention” OR “food allergy” OR “atopic dermatitis” OR “asthma”) AND TS = (infant OR newborn OR baby)*. Moreover adding the filter: *TS = (“randomized controlled trial” OR RCT OR “randomised trial”)*.

The above-mentioned searches were made with specific inclusion criteria in mind: RCTs written in English that enrolled infants at risk of developing atopic disease and in which the intervention consisted of hypoallergenic formula initiated from birth. Exclusion criteria were studies enrolling sick infants, studies in which the intervention was not initiated at birth, and non-RCTs.

Both HMV and AMS screened independently screened the records and reports retrieved. HMV collected data from each report. All results that were compatible with each outcome domain were sought i all studies.

### Risk of bias assessment

A risk of bias assessment was made using the *Cochrane Risk of Bias 2 (RoB2)* tool. All RCTs were evaluated across five domains.

### Data synthesis

Relative risks (RR) with 95% confidence intervals were extracted directly or calculated from each study. It was calculated according to atopic disease outcomes, of which AD was the primary outcome and CMPA and asthma were the secondary outcomes.

Meta-analyses were performed using *R* (version 2025.05.1+513). Forrest-plots were made to visualize the study-results and pooled effect estimates. A random-effects model was applied to account for clinical and methodological heterogeneity across studies and to estimate the pooled relative risks (RR) with corresponding 95% confidence intervals. Statistical heterogeneity was assessed using I^2^ statistics. Subgroup analysis was based on the experimental formula used (pHF, eHF, or EF) and according to the outcome (AD, CMPA, and asthma). Assessment of publication bias (Funnelplots and Egger’s test) was not feasible due to the inclusion of fewer than ten studies.

The analytical code used for this project can be acquired upon request from the corresponding author.

## Results

This systematic review and meta-analysis followed the *Preferred Reporting Items for Systematic reviews and Meta-Analyses (PRISMA)* 2020 statement, illustrated in the PRISMA flow diagram (**Figure 1**).

**Figure 1.**
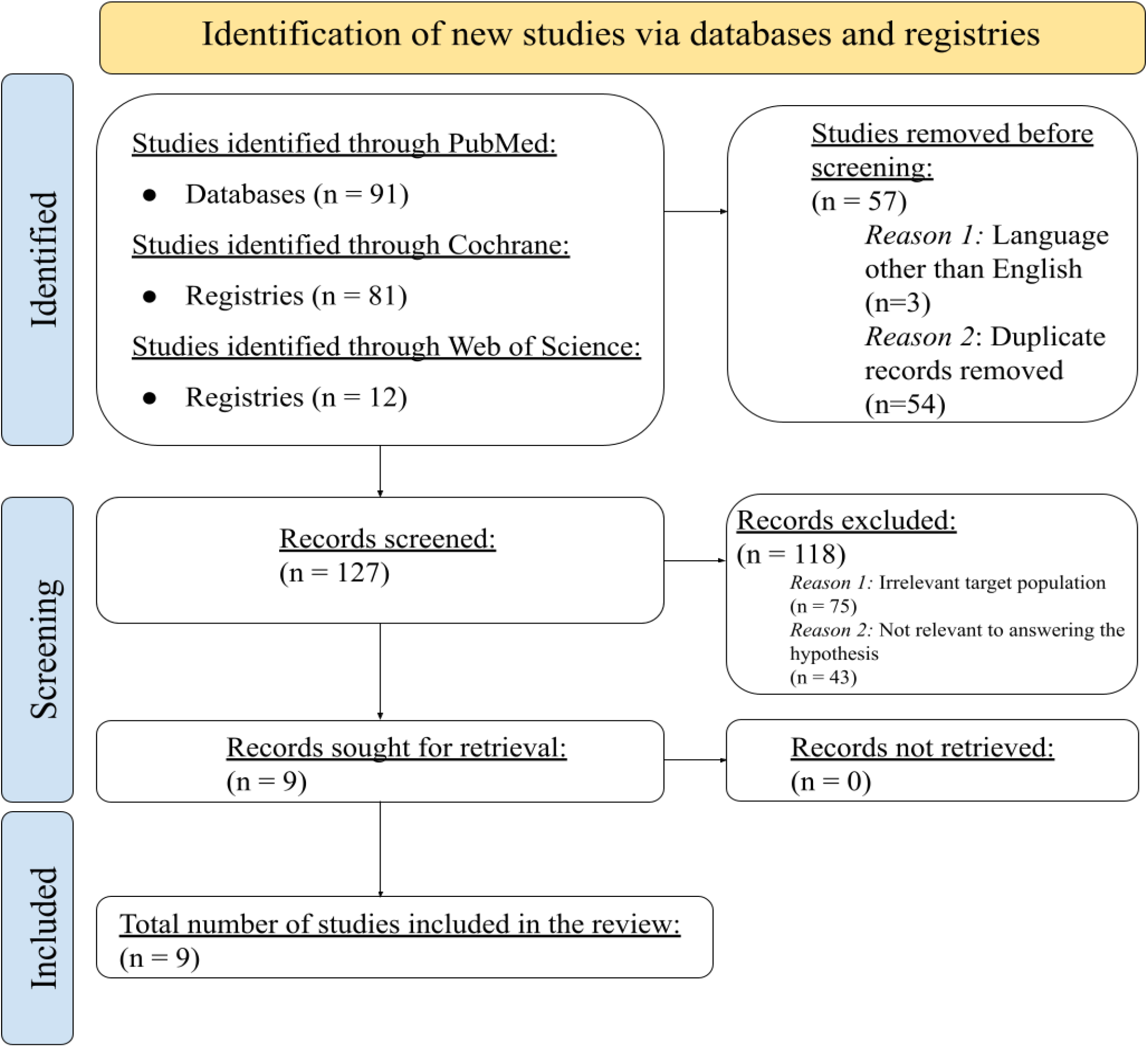
PRISMA 2020 flow diagram, showing the study selection process.

The search in Pubmed identified 91 studies, the Cochrane library identified 81 studies, and lastly, the search in Web of Science identified 12 trials.

After trials in other languages than English and duplicates were removed, 127 studies were screened. We excluded 118 records due to irrelevant target population (i.e. not infants at high risk for allergy) and due to intervention with other than formula alone (e.g. prebiotics and taste-analysis). This resulted in a total of 9 RCTs included in the final analysis. These studies are summarized in **Table 1**.

**Table 1:**
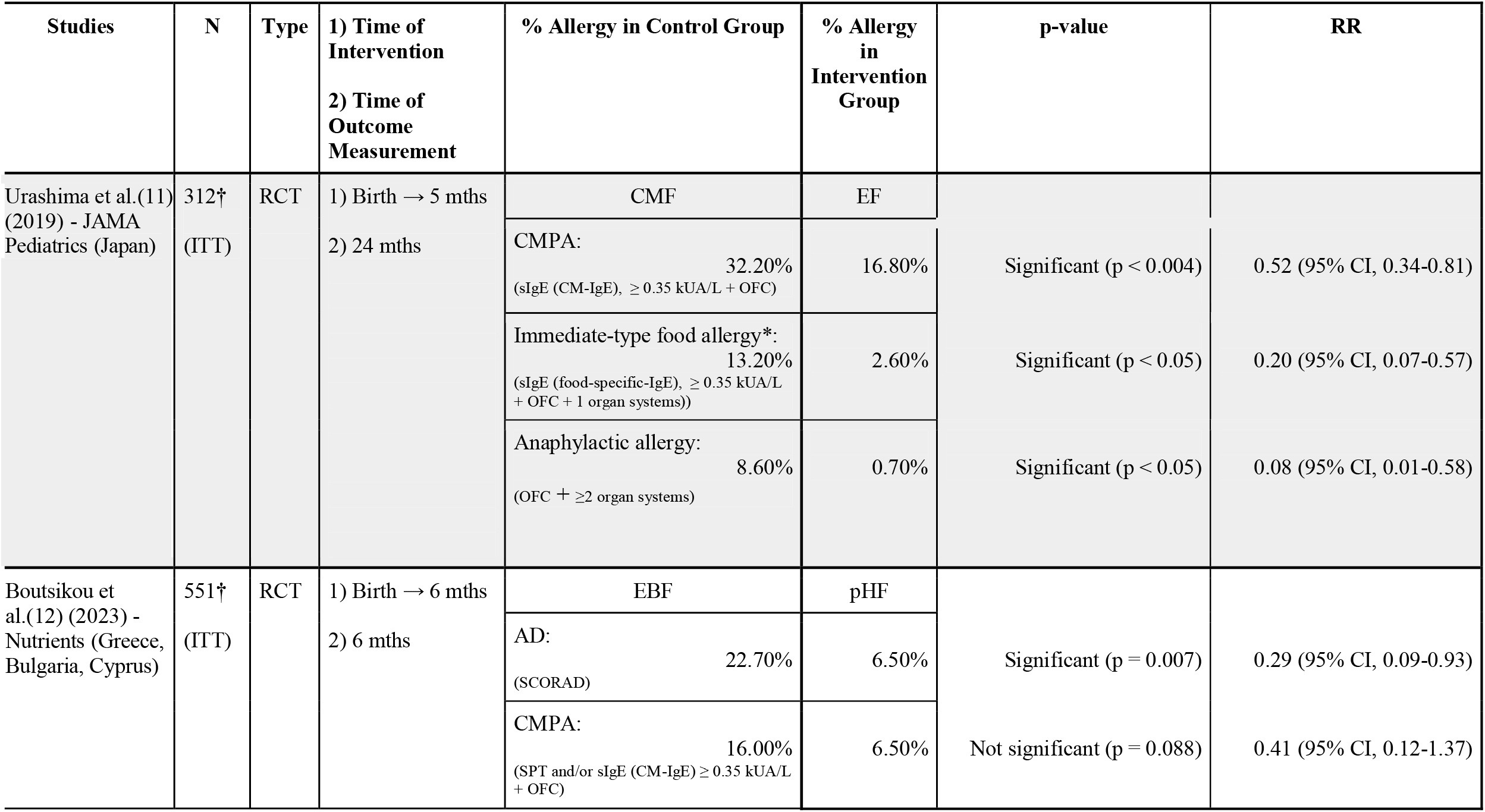
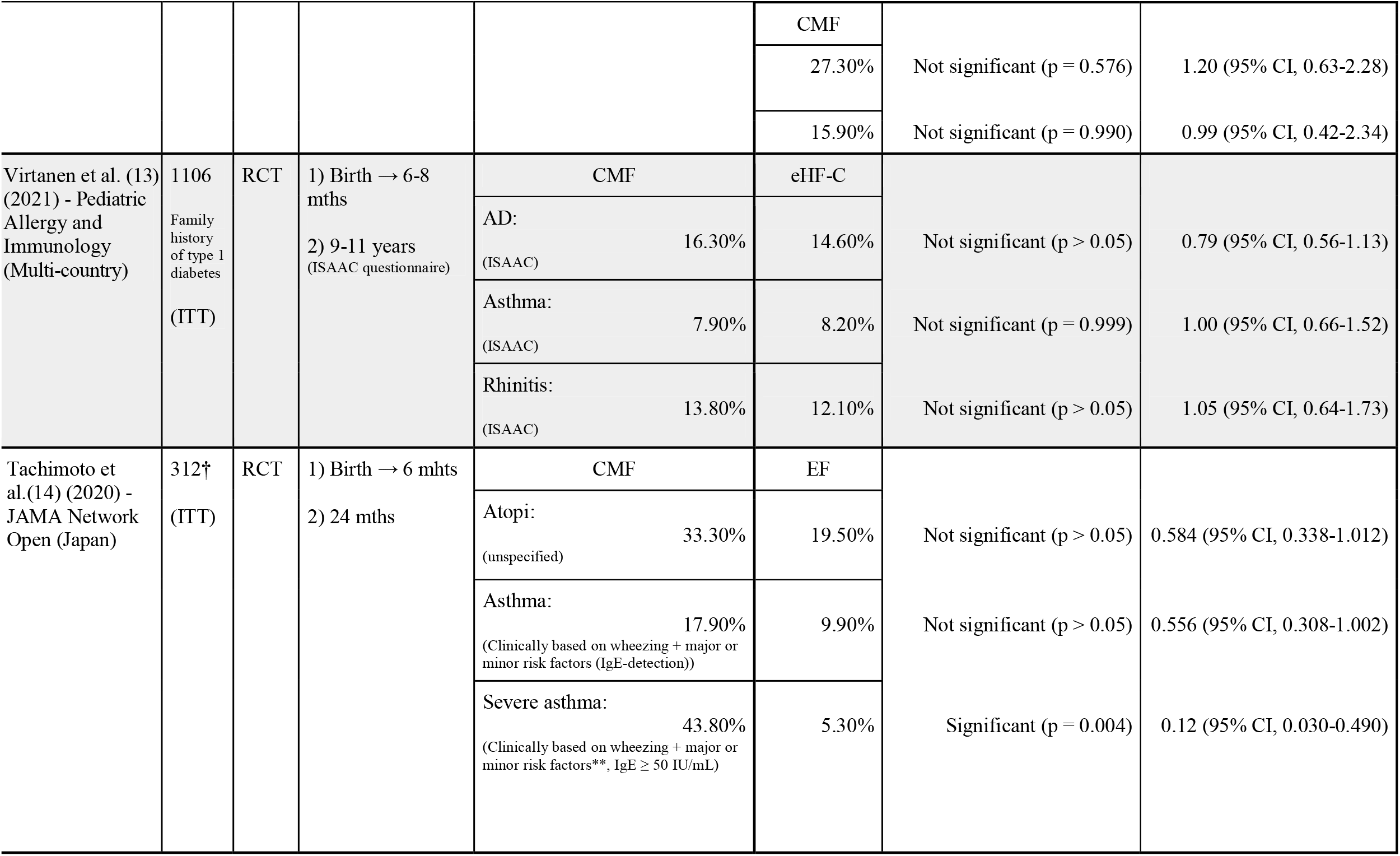
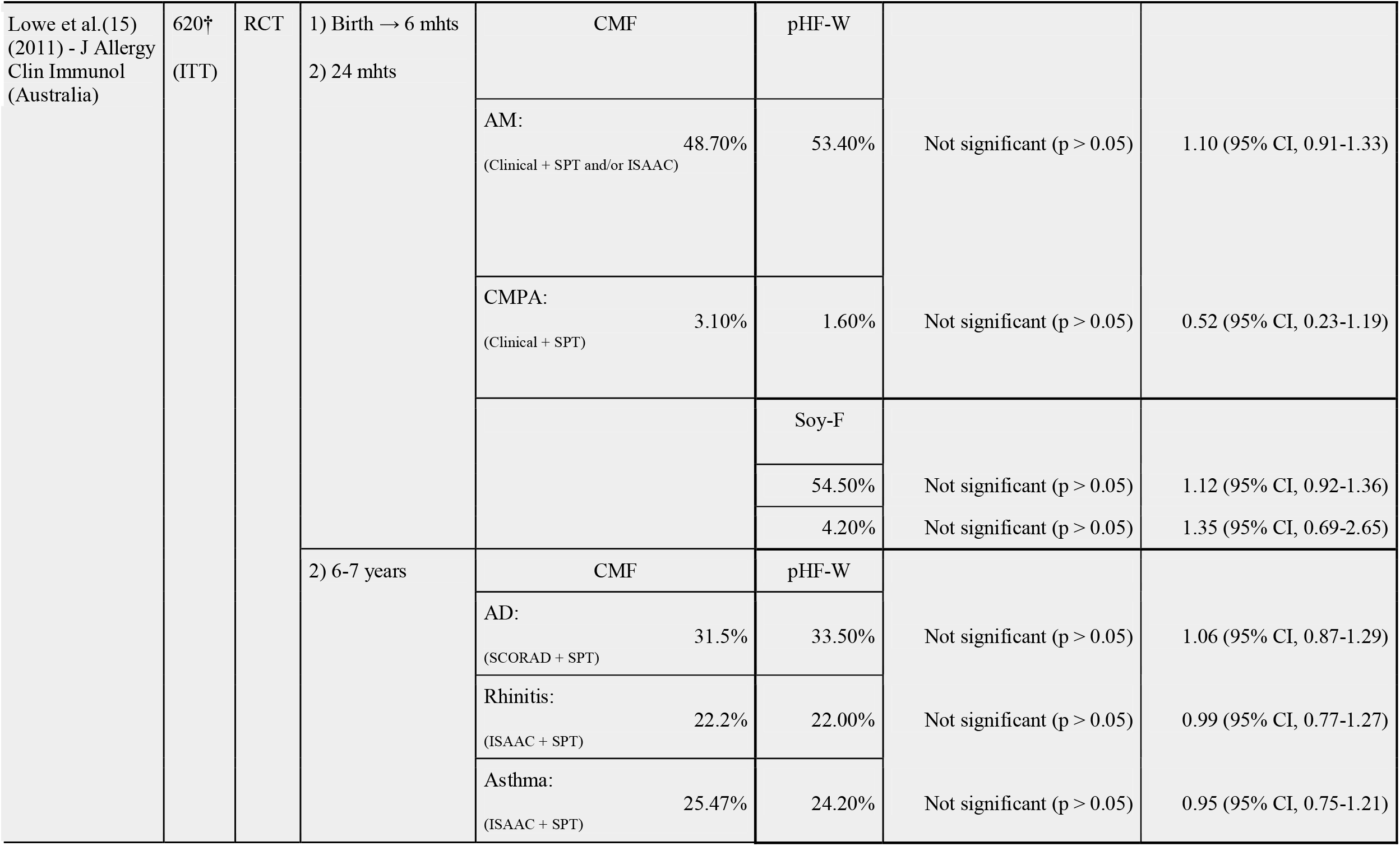
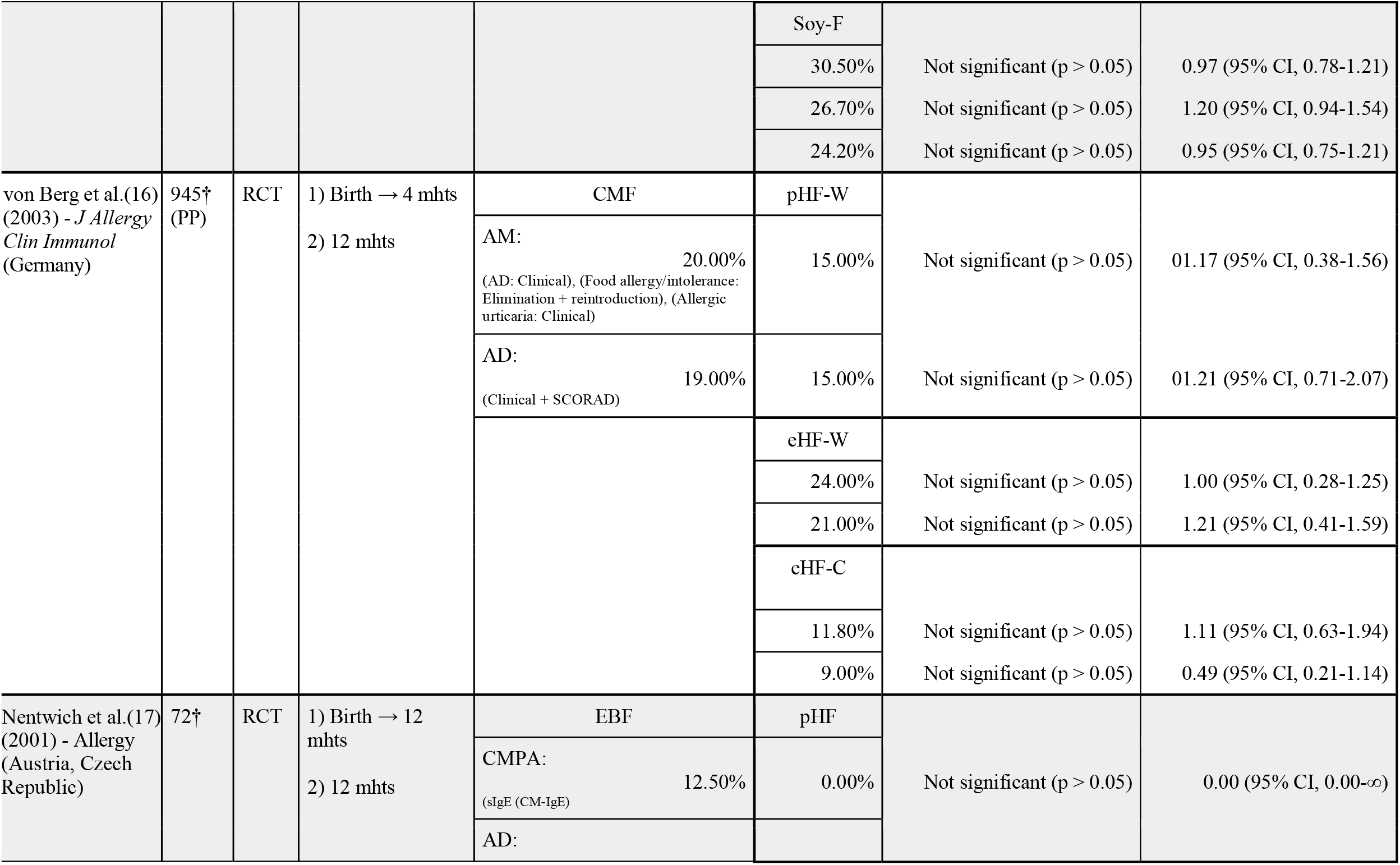
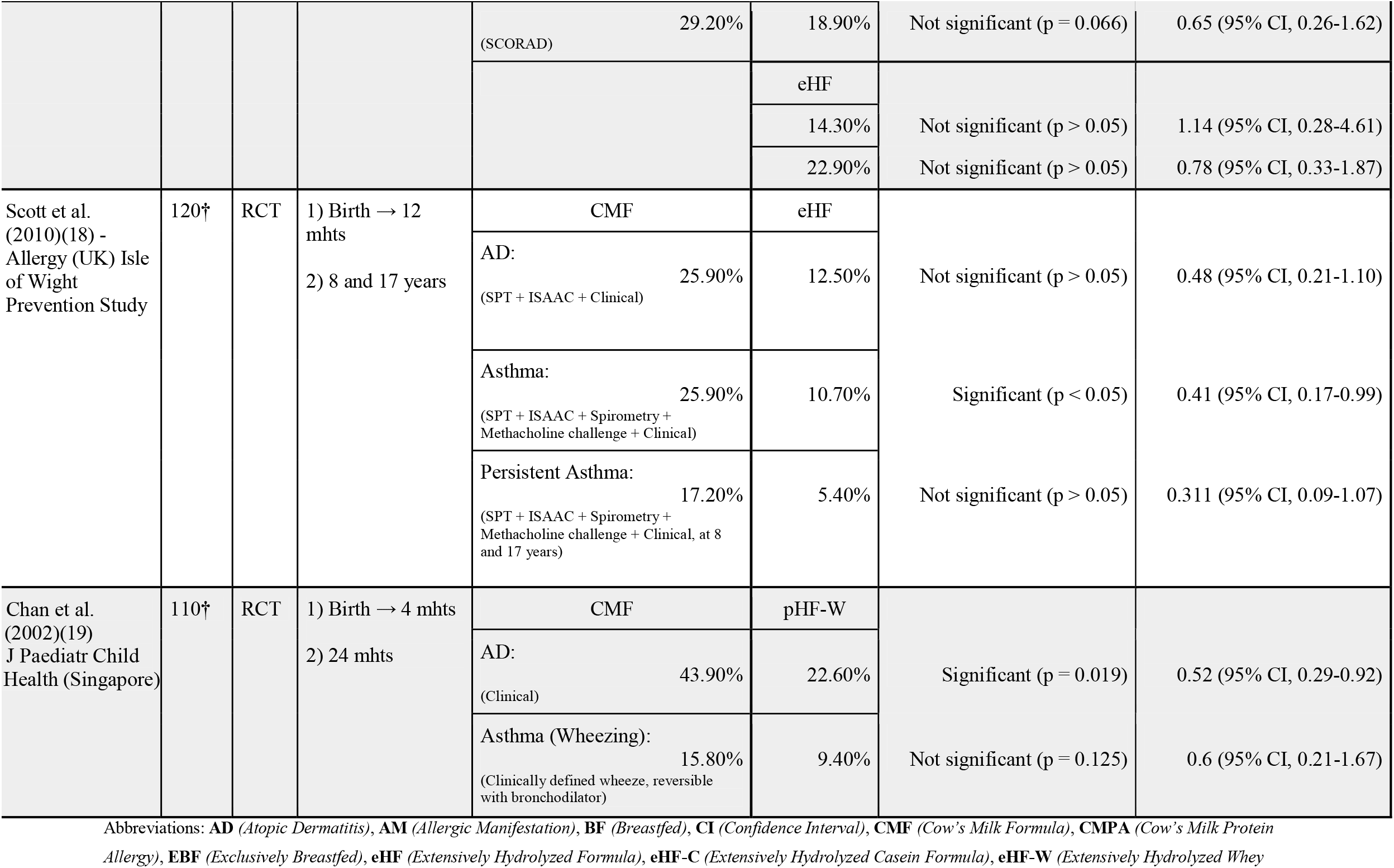
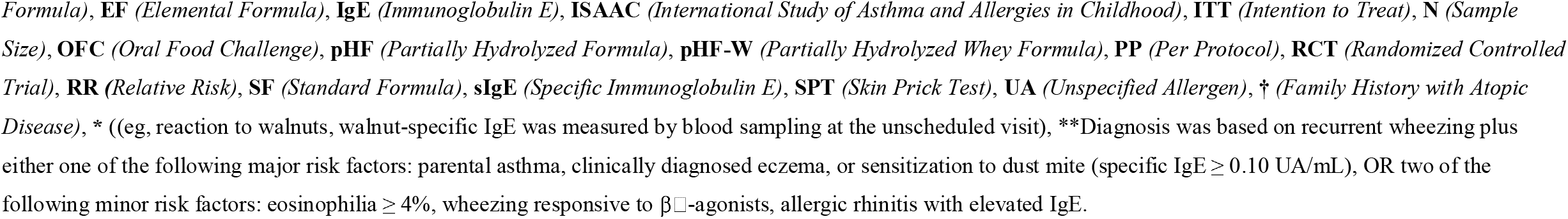
Overview of Randomized Controlled Trials (RCTs) on Hypoallergenic Infant Formula and Allergy Prevention in Children with a Family History of Atopy.

The risk of bias assessment, performed in RoB2, is visualized in **Table 2**.

**Table 2:**
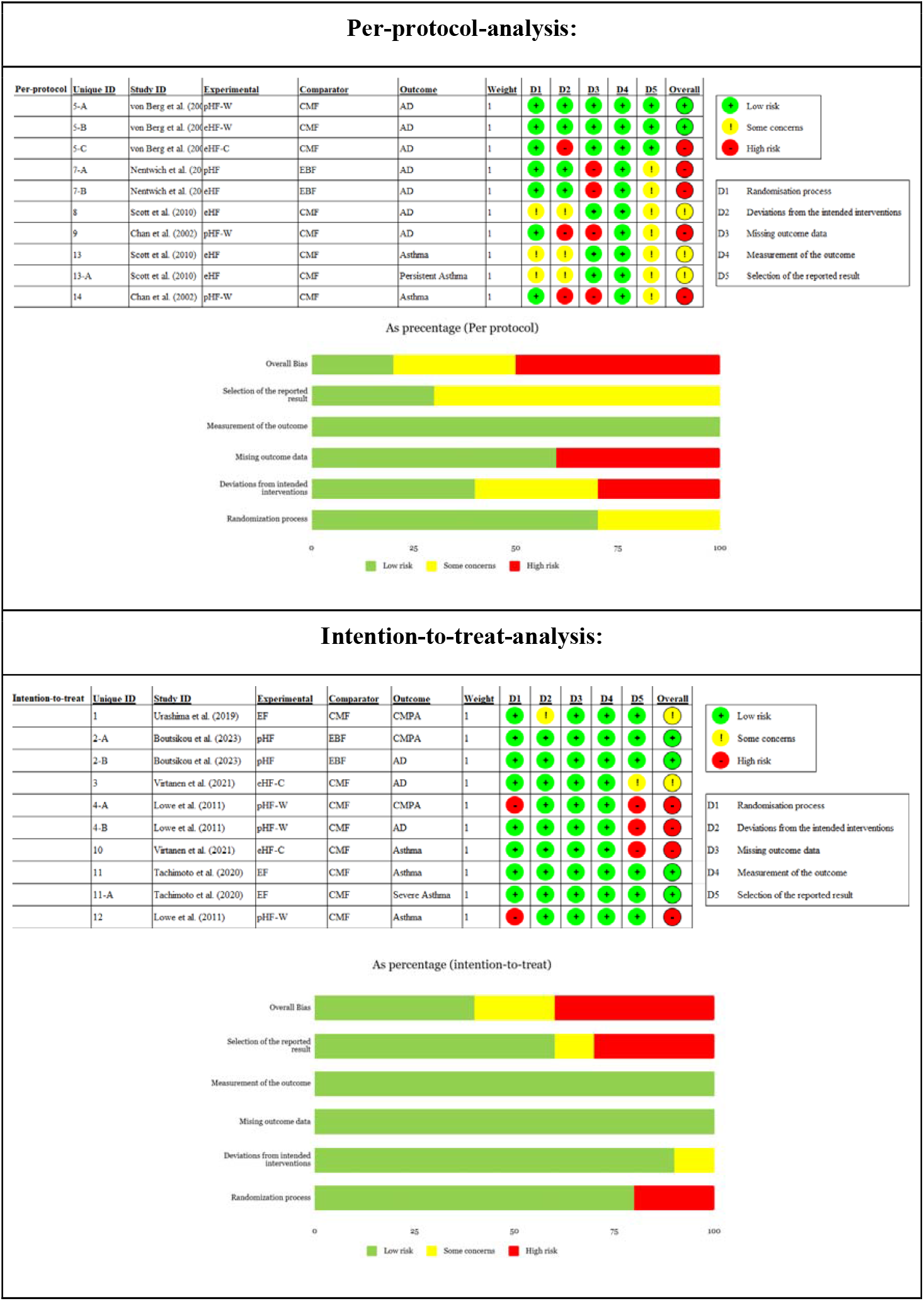
Risk of Bias Assessment.

The judgement of each domain is rated from low risk (green), some concerns (yellow) to high risk (red).

Across the studies included, many were judged as having *low risk* of bias in the majority of domains. However, in the *per protocol* (PP)-analysis, the studies *Von Berg et al*.*(16), Nentwich et al*.*(17)* and *Chan et al*.*(19)* were judged high risk due to deviations from the intended intervention and missing outcome data. In the *intention to treat* (ITT)-analysis, *Lowe et al*.*(15)* and *Virtanen et al*.*(13)* were judged as high risk due to the randomization process and selection of the reported results. Differences were noted between the PP and ITT-analysis where ITT generally had lower risk of bias compared to PP.

### Synthesis of results

Forest plots were generated for each outcome (AD, CMPA, and asthma) visualized in **Figure 2**. Pooled estimates using random-effects showed a borderline significant protective effect on hypoallergenic formula in prevention of AD (RR=0.78 [0.59-1.03], p=0.059; I^2^=46.5%). Subgroup analysis based on type of formula showed similar results **Figure 2**.

**Figure 2.**
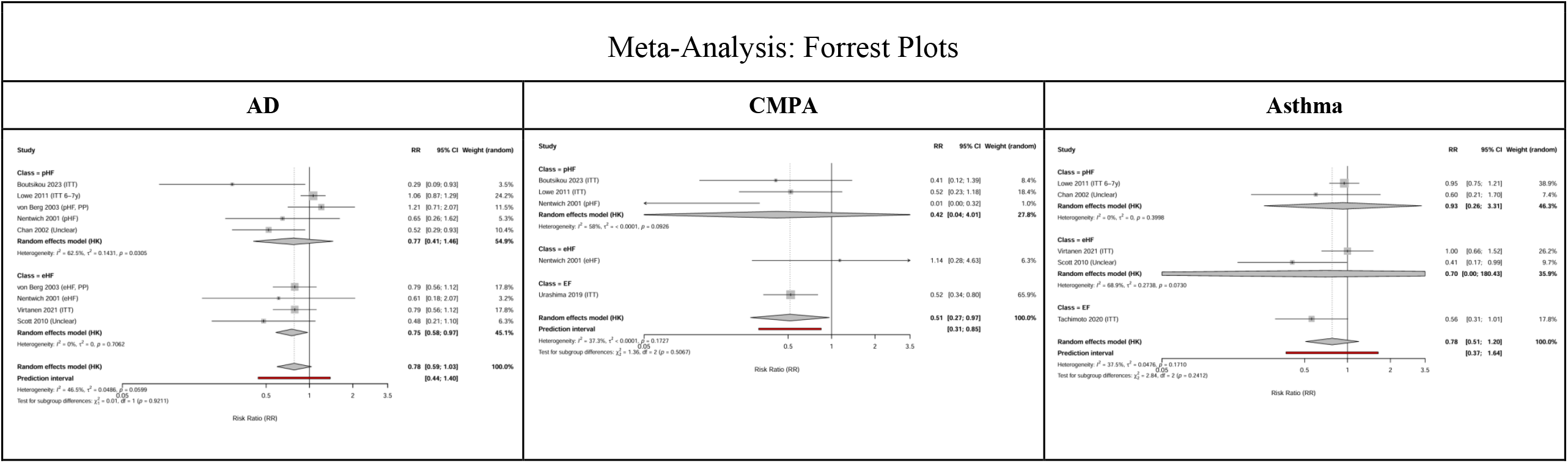
Forest plots generated in R. Subgroups (pHF, eHF, EF) are displayed under each outcome (AD, CMPA, Asthma). Note: There were not enough studies assessing EF to conduct a metaanalysis.

The pooled estimates using random-effects demonstrated a statistically significant risk-reduction in developing CMPA with the use of hypoallergenic formula (RR=0.51 [0.27-0.97], p=0.0228; I^2^=37.3%).

The pooled estimates using random-effects did not show any significant preventative effect of hypoallergenic formula in the prevention of developing asthma (RR=0.78 [0.51-1.20], p=0.171; I^2^=37.5%). However, a borderline trend towards a protective effect was observed in the eHF-subgroup (**Figure 2**).

Heterogeneity across studies was assessed using the I^2^ statistic in accordance with PRISMA guidelines. The analyses demonstrated moderate heterogeneity for AD (I^2^=46.5%) and asthma (I^2^=37.5%), and low-to-moderate heterogeneity for CMPA (I^2^=37.3%). These findings indicate some variability between trials, which is expected given differences in study design, follow-up periods, and diagnostic criteria.

## Discussion

This systematic review and meta-analysis included nine RCTs with infants at high-risk of developing atopic disease. A trend toward reduced risk of developing AD was observed, which was borderline significant and consistent across subgroups. Although the overall direction of the estimates suggested a possible trend toward risk reduction, this did not reach conventional statistical significance. The risk-reduction for developing CMPA was found significant in the assessment of hypoallergenic formula (RR=0.5, I^2^=37.3%). This finding was supported by both subgroups; however, as the EF subgroup was based on a single RTC (*Urashima et al. (2019)*), the interpretation is limited.

For asthma-prevention, a non-significant trend toward risk-reduction was observed. Subgroup results were directionally consistent across pHF, eHF and EF, although precision was limited and some estimates were driven by single trials.

Overall, the studies showed a good methodological quality. The lower bias observed in the ITT analysis refers specifically to the studies included in this review, where ITT estimates appeared more robust due to less impact from dropouts.

Strengths of this review include a comprehensive search across multiple databases, only including RCTs, risk assessment used with the tool RoB2, and the PP/ITT separation in the bias analysis. The majority of RCTs included were ITT, which generally have a smaller bias than PP. Where ITT or PP analysis was not specified, studies were assumed to be ITT. Limitations of this review include the heterogeneity across the studies in terms of study size, inclusion criteria, length of intervention and follow-up, methodology in the diagnosis, and generally the limited number of studies in each subgroup analyses. OFC is known as the strongest diagnostic method to define food allergy and is not used in all RCTs where SPT and/or sIgE-test in combination with history of reactions are sometimes used, which provide weaker evidence.

Although this meta-analysis was not designed to explore mechanisms, potential biological explanations should be considered. The hydrolyzed formula contains smaller peptides and therefore decreases antigenicity and possibly modulates immune tolerance. The lack of definitive statistically significant effect for AD and asthma may reflect the multifactorial pathogenesis of these diseases, where genetics, epigenetics, and environmental factors interact beyond early dietary influences.

The current evidence base remains limited but suggestive of a modest protective effect, particularly for CMPA. While results across studies are not entirely consistent, the overall direction of evidence supports a possible benefit in high-risk infants. Moreover, heterogeneity in study design, including differences in formula type, length of intervention, duration of follow-up, outcome definitions, diagnostic approaches, and overall study quality, further contributes to the uncertainty.

There is currently insufficient evidence to universally recommend hypoallergenic formula as a protective measure against AD or asthma, but a potential benefit cannot be excluded, especially in infants with a strong family history of atopy. In the case of CMPA, hypoallergenic formula has a possible protective effect when formula is needed in the first week of life and may be considered in high-risk infants.

Although the evidence for a protective effect is limited, no safety concerns or adverse effects were reported in the RCTs included, suggesting minimal safety concerns. The decision to use hypoallergenic formula may be guided more by cost and availability rather than safety considerations.

Disadvantages may be the costs and palatability of the hypoallergenic formula on the market today.

Future research should focus on larger, well-designed RCTs with standardized outcome-definitions and diagnoses (i.e. OFC in CMPA and Hanifin&Rajka criteria in AD), subgroup analysis based on types of hypoallergenic formula (pHF, eHF, and EF) and consideration of genetic predispositions (i.e. filaggrin status).

This could further clarify the benefit and help reduce the burden of atopic disease.

## Conclusion

This systematic review and meta-analysis, comprising nine RCTs, suggests a borderline protective effect of hypoallergenic formula for the prevention of AD. A significant risk reduction was observed for CMPA (RR ≈ 0.5). Importantly, not all CMPA diagnoses were confirmed by oral food challenge, the gold standard, which may limit the robustness of these findings. Overall, hypoallergenic formula appears to show a non-significant trend toward reduced risk of AD and asthma, while demonstrating potential efficacy in reducing the risk of CMPA in high-risk infants, suggesting potential patient-specific utility in these infants. Larger, well-designed randomized trials with standardized methodology are needed to confirm these protective effects.

## Supporting information

PRISMA checklist

## Data Availability

All data produced in the present study are available upon reasonable request to the authors

https://www.crd.york.ac.uk/PROSPERO/search

## Abbreviations

AD: Atopic Dermatitis
AM: Allergic Manifestation
BF: Breastfed
CI: Confidence Interval
CMF: Cow’s Milk Formula
CMP: Cow’s Milk Protein
CMPA: Cow’s Milk Protein Allergy
EBF: Exclusively Breastfed
eHF: Extensively Hydrolyzed Formula
eHF-C: Extensively Hydrolyzed Casein Formula
eHF-W: Extensively Hydrolyzed Whey Formula
EF: Elemental Formula
HIF: Hypoallergenic Infant Formula
ISAAC: International Study of Asthma and Allergies in Childhood
ITT: Intention to Treat
OFC: Oral Food Challenge
pHF: Partially Hydrolyzed Formula
pHF-W: Partially Hydrolyzed Whey Formula
PP: Per Protocol
RCT: Randomized Controlled Trial
RR: Relative Risk
SF: Standard Formula
sIgE: Specific Immunoglobulin E
SPT: Skin Prick Test
TS: Topic Search
UA: Unspecified Allergen

## Notes

**<u>Source of Funding:</u>** All funding received by COPSAC is listed on www.copsac.com. The Lundbeck Foundation (Grant no R16-A1694); The Ministry of Health (Grant no 903516); Danish Council for Strategic Research (Grant no 0603-00280B) and The Capital Region Research Foundation have provided core support to the COPSAC research center.

**<u>Conflict of interest:</u>** AMS: Advisory board for ALK and paid speaker for ALK, Stallergenes Greer, and ThermoFisher Scientific. All other authors declare no potential, perceived, or real conflict of interest regarding the content of this manuscript. The funding agencies did not have any role in design and conduct of the study; collection, management, and interpretation of the data; or preparation, review, or approval of the manuscript. No pharmaceutical company was involved in the study.

### Competing Interest Statement

The authors have declared no competing interest.

### Clinical Protocols

https://www.crd.york.ac.uk/PROSPERO/search

### Funding Statement

The study did not receive any direct funding. All funding received by COPSAC is listed on www.copsac.com. The Lundbeck Foundation (Grant no R16-A1694); The Ministry of Health (Grant no 903516); Danish Council for Strategic Research (Grant no 0603-00280B) and The Capital Region Research Foundation have provided core support to the COPSAC research center.

